# Matrix Metalloproteinase-9 Mediates Endothelial Glycocalyx Degradation and Correlates with Severity of Hemorrhagic Fever with Renal Syndrome

**DOI:** 10.1101/2025.02.05.25321774

**Authors:** Chloé Jacquet, Rasmus Gustafsson, Ankit Kumar Patel, Magnus Hansson, Gregory Rankin, Fouzia Bano, Julia Wigren Byström, Anders Blomberg, Johan Rasmuson, Simon Satchell, Therese Thunberg, Clas Ahlm, Marta Bally, Anne-Marie Fors Connolly

**Author notes:** Lead contact: Anne-Marie Fors Connolly. Corresponding author: Anne-Marie Fors Connolly, MD, PhD, Infectious Diseases, Dept. of Clinical, Microbiology, Building 6M 4th floor, Umeå University Hospital, 901 85 Umeå, Sweden.

## Abstract

Hemorrhagic fever with renal syndrome (HFRS) caused by Puumala virus (PUUV) leads to vascular dysfunction contributing to acute kidney injury and pulmonary complications. The endothelial glycocalyx (eGLX) is crucial for vascular integrity, and its degradation may exacerbate disease severity. In this study, we examined the association between eGLX degradation and renal and pulmonary dysfunction in 44 patients with laboratory-confirmed PUUV infection. We measured plasma levels of eGLX degradation markers—syndecan-1, heparan sulfate, soluble thrombomodulin, and albumin— and found that these correlated with severe acute kidney injury and the need for oxygen therapy. In vitro experiments showed that matrix metalloproteinase-9 (MMP-9) and heparanase can degrade eGLX components, but albumin at physiological concentrations can mitigate this degradation and protect endothelial barrier function. These findings indicate that eGLX degradation contributes to HFRS pathogenesis and suggest that targeting the eGLX could be a therapeutic strategy to improve patient outcomes.

## Introduction

Hemorrhagic fever with renal syndrome (HFRS) is a viral disease caused by orthohantaviruses, among them Puumala virus (PUUV). It is characterized by influenza-like symptoms such as fever, headache, myalgia, arthralgia, as well as renal dysfunction, respiratory symptoms, and thrombocytopenia. PUUV infection leads to endothelial damage without cytopathic effect^1^, causing vascular dysfunction that contributes to acute kidney injury and pulmonary dysfunction^2^. Our previous study was the first to demonstrate an association between endothelial glycocalyx (eGLX) degradation and the severity of HFRS^3^. The involvement of eGLX in HFRS pathogenesis has also been reported by two other studies^4,5^. Furthermore, the role of endothelial cells in HFRS is underscored by our nationwide studies in Sweden, which identified PUUV infection as an independent risk factor for acute myocardial infarction and stroke ^6^ and deep vein thrombosis and pulmonary embolism^7^.

The eGLX consists of a complex network of proteoglycans, glycosaminoglycans (GAGs) and adsorbed plasma proteins such as albumin^8^. With the exception of hyaluronan, which is attached to the cell membrane through the CD44 receptor, GAGs such as heparan sulfate (HS)—representing 50% to 90% of the eGLX GAG pool—are attached to proteoglycan cores like syndecan-1 (SDC-1) or thrombomodulin (TM)^8^. Albumin, a negatively charged plasma protein of 69 kDa^9^, plays an important role in maintaining glycocalyx integrity and physiological function^10^. The eGLX plays a critical role in maintaining the integrity and function of blood vessels by regulating permeability, inflammation, adhesion of leukocytes and platelets, and coagulation^8^. It acts as a protective barrier against the passage of macromolecules, pathogens, and inflammatory cells from the bloodstream into the underlying tissues^8^. Alteration of the eGLX allows immune cells and inflammatory mediators to access endothelial cells, leading to enhanced inflammation and tissue damage, and likely involved in the pathogenic mechanisms underlying several diseases such as acute cardiovascular events^11^, diabetes mellitus^12^, renal dysfunction^13^, sepsis^14^ and other infectious diseases^8^. Sheddases, such as matrix metalloproteinases, heparanase, chondroitinase and others, are enzymes that degrade the eGLX^15^. Notably, urinary levels of heparanase have been associated with PUUV severity, indicating a potential role of this sheddase in HFRS pathogenesis^5^.

In this study, we aimed to determine the potential association between eGLX degradation with renal and pulmonary dysfunction during HFRS, identify potential mediators of eGLX degradation and investigate whether albumin could protect against eGLX degradation.

## Methods

### Study cohort and sample collection

A total of 44 individuals with laboratory-verified PUUV infection were included in this study after providing informed written consent at Umeå University Hospital between January 2008 and June 2014. Diagnosis was based on typical clinical manifestations of HFRS and confirmed by detecting immunoglobulin G (IgG) and IgM antibodies to PUUV using an immunofluorescence assay developed in the clinic^16^. Clinical symptoms, routine laboratory tests, data on the need for oxygen supplementation, and patient samples were collected consecutively during the acute phase and at follow-up visits. Peripheral venous blood samples were collected in vacuum tubes containing sodium heparin (Becton Dickinson, Franklin Lakes, NJ, USA) and centrifuged at 1,500 g for 20 minutes. Plasma was aliquoted and immediately stored at –80 °C until analysis. Ethical approval (Dnr 07-16M) was granted by the Regional Ethical Review Board in Umeå, Sweden.

### Clinical and routine laboratory analyses

Plasma levels of creatinine, cystatin C and albumin were measured according to standard clinical routine at an accredited laboratory at the Department of Clinical Chemistry, Umeå University Hospital. Plasmatic NGAL was measured using Gentian kit on the Roche Cobas automation platform at Clinical Chemistry Department, Karolinska Institute, Stockholm, Sweden.

### Quantification of endothelial glycocalyx degradation markers in plasma

Levels of syndecan-1 (SDC-1) and soluble thrombomodulin (sTM) were measured using commercial enzyme-linked immunosorbent assay (ELISA) kits (Gen-Probe, Diaclone, France). Heparan sulfate (HS) levels were quantified using ELISA kits from Cusabio Technology LLC, USA. Active matrix metalloproteinase-9 (MMP-9) was assessed using the Sensolyte Plus 520 MMP-9 assay kit (AnaSpec, USA). All assays were performed according to the manufacturers’ instructions.

### Characterization of acute kidney injury and pulmonary dysfunction

Acute kidney injury (AKI) during HFRS was assessed according to the Kidney Disease: Improving Global Outcomes (KDIGO) 2012 guidelines^17^. Patients were categorized into “no or mild AKI” (stages 0–1) or “severe AKI” (stages 2–3). Staging was based on increases in plasma creatinine (P-creatinine, μmol/L): an increase of more than 1.5-fold from baseline indicated stage 1, 2-fold increase indicated stage 2, and 3-fold increase or P-creatinine above 353.6 μmol/L indicated stage 3. Severe AKI was defined as patients who met the criteria for AKI stage 2 or 3 at any time during HFRS.

Pulmonary dysfunction was defined by the requirement of supplemental oxygen therapy during HFRS, as determined by the treating physician at the Infectious Disease Clinic, Umeå University Hospital.

### Enzymes and recombinant proteins

Recombinant human heparanase (rhHPSE; #7570-GH-005, R&D Systems, Sweden) was diluted in rhHPSE assay buffer comprising 20 mM Tris-HCl and 4 mM CaCl₂ (pH 7.5). Recombinant human MMP-9 (rhMMP-9; ab168863, Abcam, Sweden) was diluted in rhMMP-9 assay buffer containing 50 mM Tris, 10 mM CaCl₂, 150 mM NaCl, and 0.9 mg/mL ZnCl₂ (pH 7.5). Recombinant human syndecan-1 His-tagged (rhSDC-1; #2780-SD-050, R&D Systems, USA) was reconstituted at 500 µg/mL in phosphate-buffered saline (PBS), aliquoted, and stored at –80 °C according to the manufacturer’s instructions. Heparan sulfate (IntelliHep Ltd., UK) was biotinylated following the method described by Thakar *et al.*^18^ and Bano *et al.*^19^ aliquoted, and stored at –20 °C.

### Preparation of synthetic vesicles

Synthetic vesicles were prepared using 1-palmitoyl-2-oleoyl-sn-glycero-3-phosphocholine (POPC) and 1,2-dioleoyl-sn-glycero-3-phosphoethanolamine-N-(cap biotinyl) (DOPE-Cap-Biotin) lipids (Avanti Polar Lipids Inc., USA) as described previously^19,20^. Briefly, the lipids were dissolved in chloroform at concentrations of 100 g/L (POPC) and 25 g/L (DOPE-Cap-Biotin). A lipid mixture of 95% POPC and 5% DOPE-Cap-Biotin was prepared in chloroform and dried overnight under vacuum in a round-bottom flask to form a thin lipid film. The dried lipid film was resuspended in PBS to a final concentration of 2 mg/mL. The suspension was extruded 11 times through a 50 nm polycarbonate membrane using a mini-extruder (Avanti Polar Lipids Inc., USA) to produce small unilamellar vesicles (SUVs). The SUVs were collected and stored at 4 °C until use.

### Quartz Crystal Microbalance with Dissipation Monitoring

Quartz Crystal Microbalance with Dissipation Monitoring (QCM-D) experiments were performed using an AWS X4 multichannel QCM-D system (AWSensors, Spain) equipped with 14 mm diameter silica coated SiO₂ sensors with 5 kHz fundamental frequency (AWS SNS 000049A, Spain). Prior to use, sensors were cleaned with a 2% sodium dodecyl sulfate (SDS) solution for 30 minutes, rinsed with Milli-Q water, and dried under a flow of nitrogen gas. The sensors were then activated using UV/ozone (Bioforce Nanoscience, USA) treatment for 45 minutes.

Experiments were conducted at 37 °C with a flow rate of 20 µL/min unless otherwise stated. Degassed HEPES buffer (10 mM HEPES, 150 mM NaCl, pH 7.4) filtered with 0.2 μm filters (Sarstedt, Germany) was used as the running buffer and for washing steps between injections unless otherwise specified. After establishing a stable baseline, SUVs at 100 µg/mL were injected to form a supported lipid bilayer on the sensor surface. Streptavidin (20 µg/mL) was then injected for 15 minutes. For making a pure HS film, b-HS was injected at 50µg/mL until saturation of the signal before stopping the flow for 30 min incubation in the cell chamber. To immobilize a stable recombinant human syndecan-1 His-tagged (rhSDC-1) platform, Tris-NTA-biotin (10 µg/mL, 1 mM NiCl₂; #BR1001202, Biotechne-Rabbit, Germany) was injected for 15 minutes on the streptavidin-presenting surface followed by rinsing with HEPES buffer and injection of rhSDC-1 at 50 µg/mL saturation of the signal before stopping the flow for 30 min incubation in the cell chamber.

To investigate the effect of albumin, recombinant human serum albumin (rHSA; #A9731, Merck, USA) was injected at concentrations of 40, 20, or 10 mg/mL for 30 minutes before switching to the enzyme buffer. For enzymatic treatment, rhHPSE or rhMMP-9 enzymes were injected at 500 ng/mL in their respective assay buffers, followed by incubation for 1 hour at 37 °C under stagnant conditions. After incubation, the sensor surface was washed with the enzyme buffer. The QCM-D response, frequency (Δf) and dissipation (ΔD) changes were recorded at six overtones (*i* = 3, 5, 7, 9, 11, 13) and reference value was defined as signal obtained during buffer infection before b-HS or rh-SDC-1 injection. To evaluate the impact of enzymatic treatment on b-HS or rhSDC-1, the values of Δf and ΔD signals obtained in enzyme buffer before enzyme injection were compared with values obtained in enzyme buffer after enzyme incubation. Only data from overtone 3 are shown as data from overtones *i*= 5, 7, 9, 11, 13 gave similar information. Analysis was performed using R software (R Core Team (2021).

### Cells

Conditionally immortalized human glomerular endothelial cells (CiGEnC)^21^, kindly provided by Prof. Simon Satchell (University of Bristol, UK), were cultured at 33 °C with 5% CO₂ in EBM™-2 basal medium (#CC-3156, Lonza, Sweden) supplemented with EGM™-2 MV Microvascular Endothelial Cell Growth Medium SingleQuots™ (#CC-4147, Lonza, Sweden). Five days prior to the experimental assays, CiGEnCs were transferred to 37 °C to halt proliferation and promote the formation of tight junctions of the endothelial barrier.

### Impedance measurement for endothelial barrier integrity monitoring

The integrity of the endothelial barrier in CiGEnC cells was assessed using an Electric Cell-substrate Impedance Sensing (ECIS) system with 8-well electrode array slides (8W10E PET; Applied Biophysics, USA). Slides were coated for 1 hour at 37 °C with 50 µg/mL bovine fibronectin (Gibco™, USA) diluted in phosphate-buffered saline (PBS), followed by stabilization of the gold electrodes with 10 µM L-cysteine (#C7352, Sigma-Aldrich, USA) in Milli-Q water for 30 minutes at 37 °C. Impedance measurements at 4,000 Hz were initially performed in complete medium to establish a stable baseline over 30 minutes to 1 hour. Cells were then seeded at 2.5 × 10⁵ cells/mL at 33 °C CiGEnCs. Upon reaching confluency, CiGEnCs were shifted to 37 °C and incubated for an additional 5 days before treatment to facilitate barrier formation. Tumor necrosis factor-alpha (TNF-α; 50 ng/mL; #10291-TA, R&D Systems, USA) was used as a positive control for barrier disruption (supplementary figure 3). For experimental treatments, recombinant human heparanase (rhHPSE) or recombinant human MMP-9 (rhMMP-9) were added at 250 ng/mL in an equal mixture of media without FBS and the corresponding enzyme buffer, either alone or with recombinant human serum albumin (rHSA) at concentrations of 20 mg/mL or 40 mg/mL. Cells were incubated with these treatments for 12 hours, and impedance was continuously monitored.

### Staining for confocal microscopy

Cells were treated with rhHPSE at 250 ng/mL for 1 hour at 37 °C or with rhMMP-9 at 500 ng/mL for 12 hours in a 1:1 (v/v) mixture of cell media and the respective enzyme assay buffer. For albumin conditions, rHSA was included at 10 mg/mL, 20 mg/mL, or 40 mg/mL in the enzyme mixture. Post-treatment, cells were washed with cold PBS and fixed with 4% paraformaldehyde for 15 minutes at room temperature. Blocking was performed with PBS containing 5% milk for 1 hour at room temperature with gentle shaking. Heparan sulfate (HS) and syndecan-1 (SDC-1) were detected using mouse anti-HS IgM antibody (#370255-S, AmsBio, USA) and mouse anti-CD138 (SDC-1) IgG antibody (#NBP2-37282SS, Novus Biologicals, Sweden), respectively, at a dilution of 1:500 in PBS overnight at 4 °C. After washing with PBS, cells were incubated with goat anti-mouse IgG/IgM Alexa Fluor™ 488-conjugated secondary antibody (#10138324, Invitrogen, Fisher Scientific, Sweden) at a 1:1,000 dilution in PBS containing 5% milk for 1 hour at 37 °C. Nuclei were counterstained with DAPI (1:5,000 dilution from a 5 mg/mL stock solution; Invitrogen, Sweden) for 5 minutes. Following final washes, cells were mounted with appropriate mounting medium, and coverslips were applied. Confocal images were acquired using a Leica TCS SP8 MP multiphoton microscope.

### Statistics

Longitudinal changes in plasma and urine concentrations of eGLX degradation markers were analyzed using the generalized estimating equations (GEE) method with an exchangeable correlation structure between consecutive observations. Estimated means and standard errors were calculated and means within specified time groups were compared to those at follow-up (>30 days post-disease onset [DPDO]). The association between eGLX degradation markers and kidney damage markers (plasma cystatin C and creatinine levels) was assessed using GEE for all samples within 30 DPDO, with kidney damage markers as dependent variables. All analyses were adjusted for sex and age. Receiver operating characteristic (ROC) curve analysis was performed to determine the area under the curve (AUC) for the association between peak values of eGLX degradation markers and disease outcomes. For significant AUCs, cut-off values of eGLX degradation markers with the highest sensitivity and specificity were identified. Statistical significance was set at p < 0.05. All statistical analyses were conducted using SPSS version 25 (IBM).

## Results

### Characteristics of HFRS patients

A total of 44 patients with HFRS were included in the study; their characteristics are presented in Table 1. The majority of patients (82%) required hospitalization, with an average hospital stay of 6 days. According to the KDIGO 2012 guidelines^17^, 61% of the patients were classified as having severe AKI (stages 2–3). Oxygen supplementation was required for 12 patients (27%), as determined by the treating physicians. The levels of eGLX, sheddase and neutrophil activation (NGAL) markers were significantly increased during the acute phase compared to the follow up (>30 DPDO), apart from HPSE (supplementary Figure S1). Since HPSE levels were not increased (supplementary Figure S1G), this biomarker was not included in the remaining analyses of patient samples.

**Table 1:**
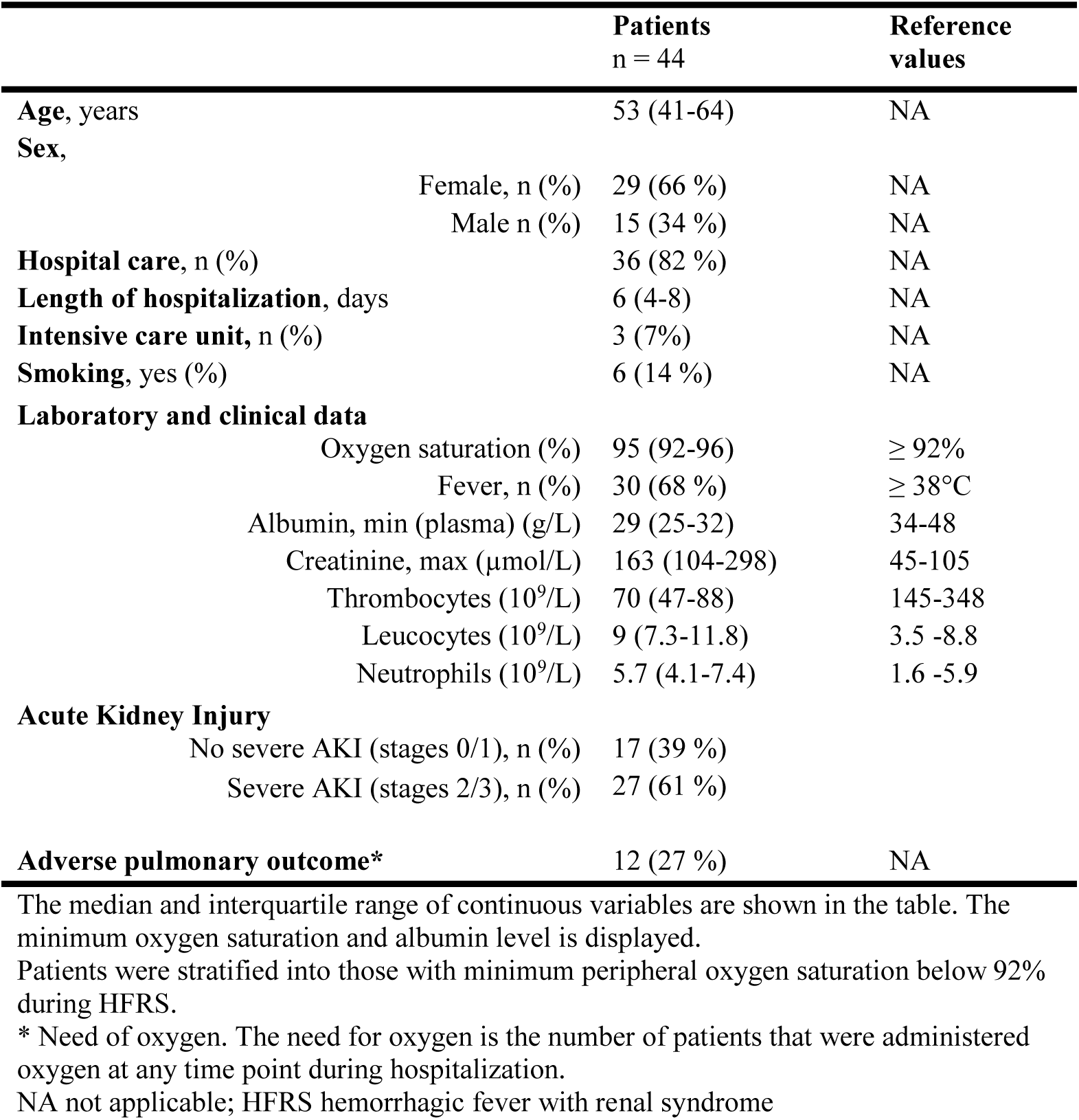
Characteristics of hemorrhagic fever with renal syndrome patients.

|  | **Patients**<br>n = 44 | **Reference**<br>**values** |
| --- | --- | --- |
| **Age, years** | 53 (41-64) | NA |
| **Sex,** |  |  |
| Female, n (%) | 29 (66 %) | NA |
| Male n (%) | 15 (34 %) | NA |
| **Hospital care, n (%)** | 36 (82 %) | NA |
| **Length of hospitalization, days** | 6 (4-8) | NA |
| **Intensive care unit, n (%)** | 3 (7%) | NA |
| **Smoking, yes (%)** | 6 (14 %) | NA |
| **Laboratory and clinical data** |  |  |
| Oxygen saturation (%) | 95 (92-96) | ≥ 92% |
| Fever, n (%) | 30 (68 %) | ≥ 38°C |
| Albumin, min (plasma) (g/L) | 29 (25-32) | 34-48 |
| Creatinine, max (μmol/L) | 163 (104-298) | 45-105 |
| Thrombocytes (10<sup>9</sup>/L) | 70 (47-88) | 145-348 |
| Leucocytes (10<sup>9</sup>/L) | 9 (7.3-11.8) | 3.5 -8.8 |
| Neutrophils (10<sup>9</sup>/L) | 5.7 (4.1-7.4) | 1.6 -5.9 |
| **Acute Kidney Injury** |  |  |
| No severe AKI (stages 0/1), n (%) | 17 (39 %) |  |
| Severe AKI (stages 2/3), n (%) | 27 (61 %) |  |
| **Adverse pulmonary outcome\*** | 12 (27 %) | NA |

The median and interquartile range of continuous variables are shown in the table. The minimum oxygen saturation and albumin level is displayed.
Patients were stratified into those with minimum peripheral oxygen saturation below 92% during HFRS.
\* Need of oxygen. The need for oxygen is the number of patients that were administered oxygen at any time point during hospitalization.
NA not applicable; HFRS hemorrhagic fever with renal syndrome

### Endothelial glycocalyx degradation associates with renal and pulmonary dysfuntion during HFRS

Patients were stratified based on clinical outcomes into two categories for renal dysfunction: (1) no AKI (stage 0) or mild AKI (stage 1), and (2) severe AKI (stages 2–3), according to KDIGO 2012 guidelines^17^. For pulmonary dysfunction, patients were categorized into (1) no requirement for supplemental oxygen, and (2) requirement for supplemental oxygen. We detected significantly different levels of eGLX markers in plasma when relating results to level of kidney dysfunction: higher levels of SDC-1, HS, sTM, and lower levels of albumin were detected in samples from patients with severe AKI, compared to patients without severe AKI (Figure 1A–D). Similarly, patients requiring oxygen supplementation exhibited significantly different levels of eGLX markers compared to those not requiring oxygen, except for SDC-1, which showed higher levels but did not reach statistical significance (Figure 1A–D). Overall, levels of the eGLX sheddase MMP-9 were higher in HFRS patients with both renal and pulmonary dysfunction (Figure 1D).

**Figure 1:**
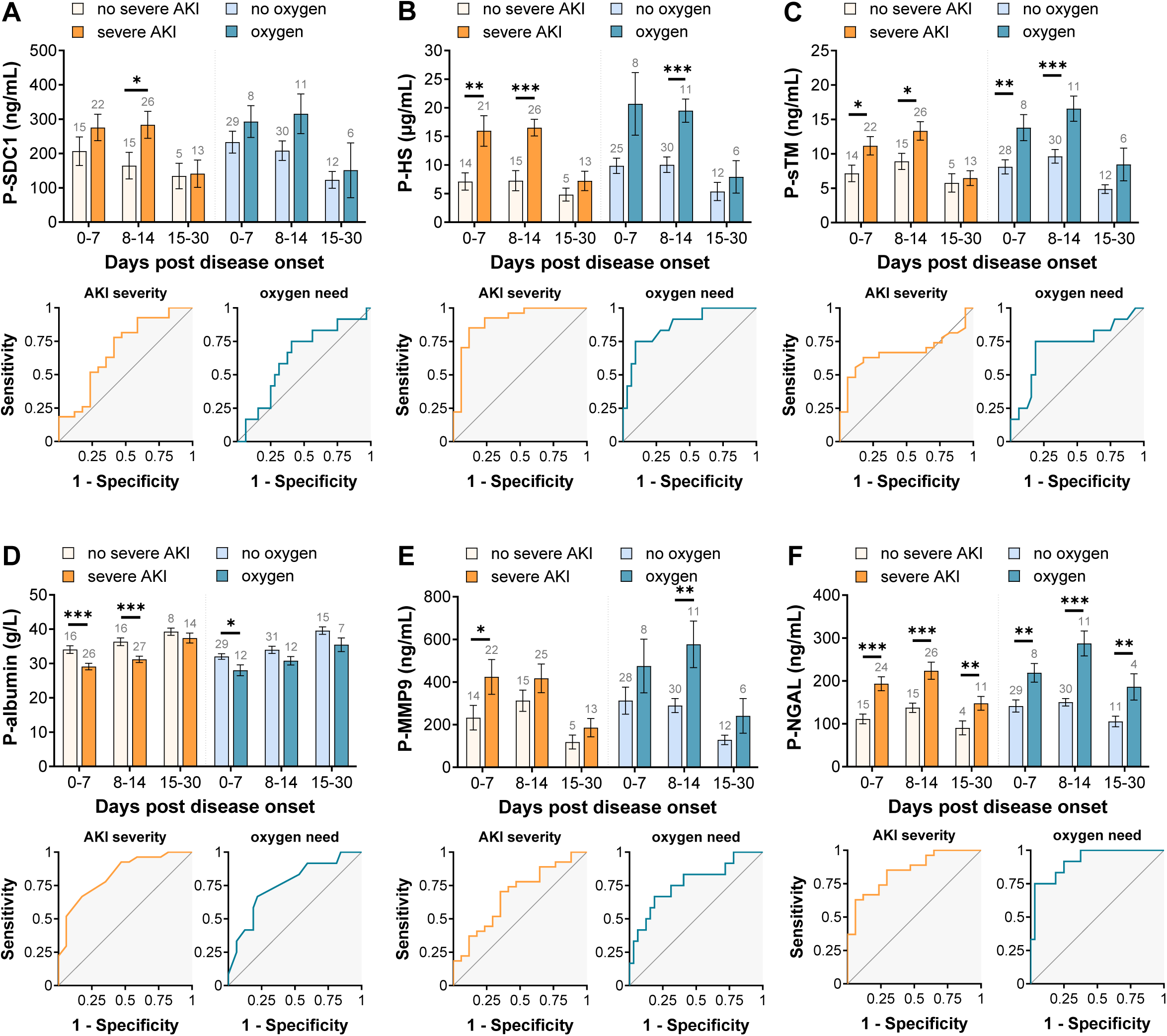
Endothelial glycocalyx degradation in HFRS patients in plasma and association with acute severe injury or need of oxygen. HFRS patients were stratified into two groups of severe AKI (stage 2/3) or not (AKI stage 0/1) according to KDIGO criteria and in two groups need of oxygen or not. The estimated mean and standard error for each group were calculated using generalized estimating equation (GEE) and adjusted for sex and age. Receiver operating characteristics curve (ROC) analysis were performed using maximum values of eGLX markers and minimal values of albumin. The timeline kinetics and ROC curves are shown for syndecan-1 **(A)**, heparan sulfate **(B)**, soluble thrombomodulin **(C)**, albumin **(D)** MMP-9 **(E)** and NGAL **(F).** Significant differences within the same time point between marker levels are indicated by asterisks (*** p < 0.001; **p < 0.01; * p < 0.05). AKI acute kidney injury; HFRS hemorrhagic fever with renal syndrome; HS heparan sulfate; KDIGO Kidney Disease Improval Global Outcomes; SDC-1 syndecan-1; P plasma; sTM soluble thrombomodulin; MMP9 matrix metalloproteinase 9 ; NGAL neutrophil gelatinase-associated lipocalin.

To determine cut-off values for eGLX biomarkers associated with renal and pulmonary dysfunction, receiver operating characteristic (ROC) curve analyses were performed using the peak values of each biomarker (minimum values for albumin) (Figure 1 and Supplementary Table 1). Shedding of eGLX biomarkers was strongly associated with renal dysfunction (except for plasma sTM) and pulmonary dysfunction (except for plasma SDC-1). Among the biomarkers, heparan sulfate (HS) demonstrated the highest area under the curve (AUC) for renal dysfunction (AUC = 0.90; 95% CI, 0.80–1.00) with a cut-off value of 10.5 μg/mL, yielding a sensitivity of 85% and specificity of 82%. HS was also strongly associated with pulmonary dysfunction (AUC = 0.87; 95% CI, 0.75–0.99) with a cut-off value of 17.9 μg/mL, sensitivity of 83%, and specificity of 72%. Additionally, the eGLX sheddase MMP-9 was significantly associated with both renal and pulmonary dysfunction.

### Endothelial glycocalyx degradation associates with MMP-9 sheddases during HFRS

Given that MMP-9 activity can lead to cleavage of SDC-1, we evaluated the association between plasma levels of MMP-9 and eGLX components during HFRS (Table 2). All eGLX biomarkers—SDC-1, HS, sTM, and albumin—showed strong associations with plasma MMP-9 levels during the disease period (<30 days post-disease onset). Furthermore, eGLX biomarkers were strongly associated with neutrophil gelatinase-associated lipocalin (NGAL), a protein known to stabilize MMP-9 and promote its activity.

**Table 2.** Association between MMP9 and NGAL, and plasma markers for endothelial surface layer degradation during HFRS.

|  | MMP-9 (ng/ml) | NGAL (ng/ml) |
| --- | --- | --- |
| **Syndecan-1 (ng/ml)** | **$\beta = 0.116$**<br>**$p < 0.001$** | **$\beta = 0.766$**<br>**$p = 0.003$** |
| **Soluble thrombomodulin (pg/ml)** | **$\beta = 3.716$**<br>**$p = 0.001$** | **$\beta = 29.57$**<br>**$p < 0.001$** |
| **Heparan sulfate (ng/ml)** | **$\beta = 9.734$**<br>**$p < 0.001$** | **$\beta = 50.986$**<br>**$p < 0.001$** |
| **Albumin (mg/l)** | **$\beta = -6.157$**<br>**$p < 0.001$** | **$\beta = -13.845$**<br>**$p = 0.045$** |

The estimated $\beta$ -coefficients from the generalized estimating equation method is shown with the corresponding P value. The $\beta$ -coefficient is the change in dependent covariate (endothelial surface layer degradation markers) following 1 unit increase for the continuous covariate MMP9. Associations shown in bold are significant. All markers were adjusted for age and sex. Only time points within 30 days post disease onset are included. MMP-9, matrix metalloproteinase-9. NGAL, neutrophil gelatinase-associated lipocalin, HFRS hemorrhagic fever with renal syndrome

### Sheddases mediate in vitro degradation of HS and SDC-1

Building on our patient cohort results, we investigated the mechanisms underlying the degradation of eGLX biomarkers, specifically focusing on SDC-1 and HS, using QCM-D. B-HS and His-tagged recombinant rhSDC-1 were attached to synthetic phospholipid membranes, enabling real-time monitoring of eGLX component degradation induced by MMP-9 or heparanase (HPSE) (Figures 2A-B). Two parameters were measured: Δf (frequency), related to layer mass and thickness, where a negative Δf value is associated with adsorption at the sensor, and ΔD (dissipation), related to the viscoelasticity of the layer^22^. After treatment with HPSE, the -Δf signal for b-HS (in enzyme buffer) was decreased by 40.8% (±7.9%), indicating substantial degradation of the amount of b-HS absorbed (Figures 2C and 2E). MMP-9 exhibited a high capacity to cleave rhSDC-1, resulting in 69.3% (±17.6%) reduction in the amount of material adsorbed (Figures 2D and 2F). In all cases, the ΔD signal decreased upon enzymatic treatment (Figures 2C-F), indicating that as the macromolecules are degraded, the adlayer became more rigid. Degradation of b-HS or rhSDC-1 is due to the activity of HSPE or MMP-9 respectively as the enzymes are unable to degrade other biotinylated or his-tagged proteins (supplementary figure 2).

**Figure 2:**
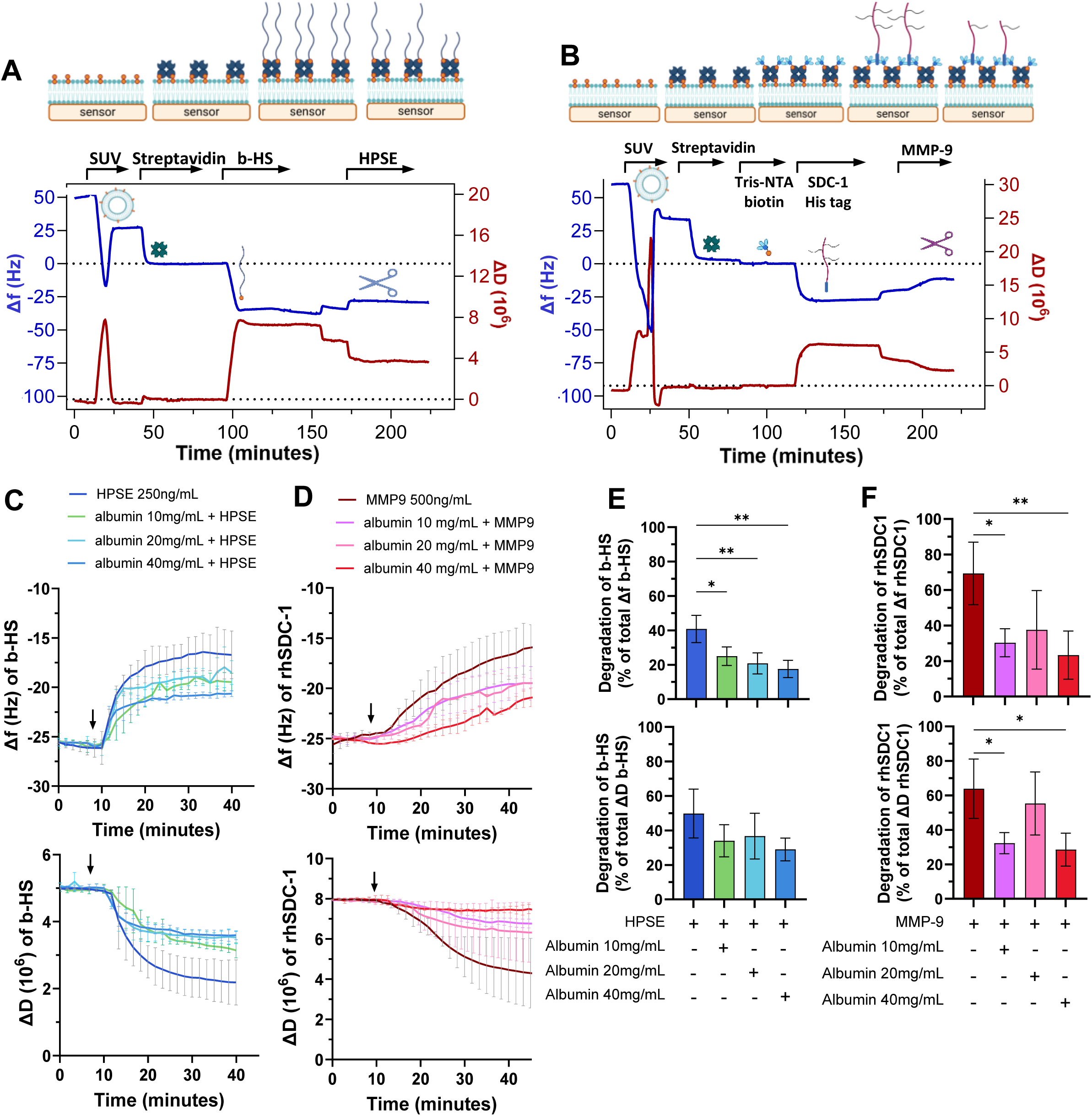
*In vitro* glycocalyx degradation mediated by heparanase and MMP-9 followed by QCM-D. **(A)** b-HS and **(B)** rhSDC-1 protein were absorbed on synthetic membrane composed by POPC-DOPE (95:5) lipids and streptavidin, plus Tris-NTA linker for rhSDC-1. After incubation with albumin at 40, 20 or 10 mg/mL, real-time degradation of b-HS by HPSE at 250 ng/mL (**C**) and rhSDC1 by MMP-9 at 500 ng/mL (**D**). Arrow indicate when the enzyme is injected. Shift in frequency (Δf) and dissipation (ΔD) due to enzyme activity was measured after enzyme activity in enzyme buffer. Percentage of Δf and ΔD signal reduction for b-HS (**E**) and rhSDC1 (**F**) after enzymatic treatment (n = 4) were calculated from the b-HS or rhSDC-1 signal in enzyme buffer (corresponding to 100%). Significant differences are indicated by asterisks (*** p < 0.001; **p < 0.01; * p < 0.05). QCM-D quartz crystal microbalance with dissipation; b-HS biotinylated heparan sulfate; rhSDC-1 recombinant human syndecan-1; MMP-9 matrix metalloproteinase 9; HPSE human heparanase.

### Albumin protects eGLX against degradation by heparanase

Given the crucial role of albumin in maintaining eGLX homeostasis, we evaluated its ability to prevent rhSDC-1 and b-HS degradation by pre-incubating them with recombinant human serum albumin before introducing the sheddases. Albumin at 40 mg/mL (normal plasma concentration) reduced eGLX degradation. In this case, the -Δf signal decreased by only 17.6% (±5%) instead of 40.8% (±7.9%) without albumin. (Figure 2C and 2E, -ΔF signal). Even at lower concentrations (20 mg/mL and 10 mg/mL), albumin showed a protective effect against HPSE activity (Figures 2C and 2E). Similarly, albumin at 40 mg/mL prevented rhSDC-1 degradation, resulting in a -Δf decrease for rdSDC-1 of 23.3% (±13.6%) instead of 69.3% (±17.6%) without albumin (Figures 2D and 2F).

### Albumin attenuates MMP-9-induced endothelial barrier disruption

To assess the impact of albumin on the eGLX layer on live cells, we incubated human glomerular endothelial cells (CiGEnCs) with increasing concentrations of albumin in the presence of MMP-9 or HPSE. Immunostaining revealed decreased SDC-1 and HS levels at the cell surface following MMP-9 and HPSE treatment, whereas the presence of albumin significantly reduced degradation of these eGLX components (Figures 3A–D). To further investigate the role of albumin, we conducted impedance assays. Unlike HPSE, MMP-9 significantly reduced endothelial barrier resistance (Figures 3E–F). Notably, albumin at 40 mg/mL effectively prevented MMP-9–induced endothelial barrier disruption.

**Figure 3:**
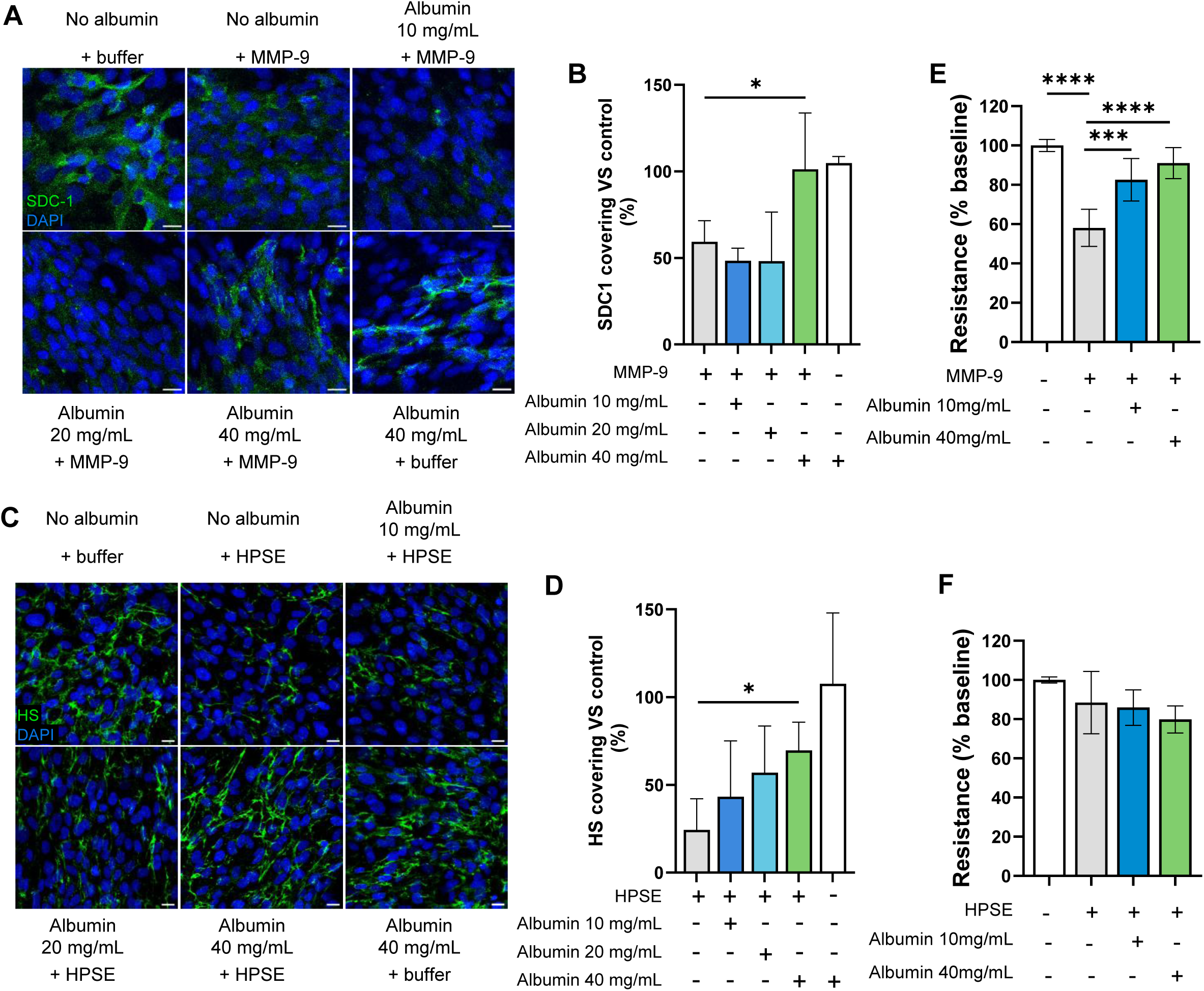
*In vitro* endothelial glycocalyx degradation and endothelial barrier disruption followed by impedance measurement. **(A)** SDC-1 staining on ciGEnC cells after incubation with MMP-9 enzyme and increased concentration of albumin. (**B**) SDC-1 fluorescence quantification after treatment compared to control (no treatment) (averaged values of 3 sections per condition, n = 3 assays). **(C)** HS staining on ciGEnC cells after incubation with HSPE enzyme and increased concentration of albumin. (**D**) HS fluorescence quantification after treatment compared to control (no treatment) (averaged values of 3 sections per condition, n = 3 assays). **(E)** Endothelial barrier resistance followed by impedance measurement (ECIS, APBiophysic) on ciGEnC after treatment with MMP-9 and albumin (averaged of 2 values per conditions, n =3 assays). **(F)** Endothelial barrier resistance followed by impedance measurement (ECIS, APBiophysic) on ciGEnC after treatment with HSPE and albumin (averaged of 2 values per conditions, n =3 assays). Significant differences are indicated by asterisks (*** p < 0.001; **p < 0.01; * p < 0.05). HS heparan sulfate ; SDC-1 syndecan-1; MMP-9 matrix metalloproteinase 9; HPSE human heparanase.

### Elevated immune cell levels associate with severe AKI and pulmonary dysfunction during HFRS

Consistent with previous analyses in our study, patients were stratified according to AKI severity stages and pulmonary dysfunction status. Leukocyte counts were significantly elevated and associated with severe AKI and the need for supplemental oxygen during HFRS (Figure 4A). Among leukocyte subtypes, lymphocyte levels increased significantly during the late stage of HFRS, while monocyte levels were markedly higher in patients with severe AKI and those requiring oxygen supplementation (Figures 4B–C). Neutrophil counts correlated with AKI and oxygen requirement during the early acute phase of disease (Figure 4D), whereas there were no significant differences in basophil levels for HFRS patients with AKI or need of oxygen (Figure 4E). Eosinophil levels were higher during the middle phase of HFRS in those with severe AKI, but with no significant differences for HFRS patients that required oxygen (Figure 4F).

**Figure 4:**
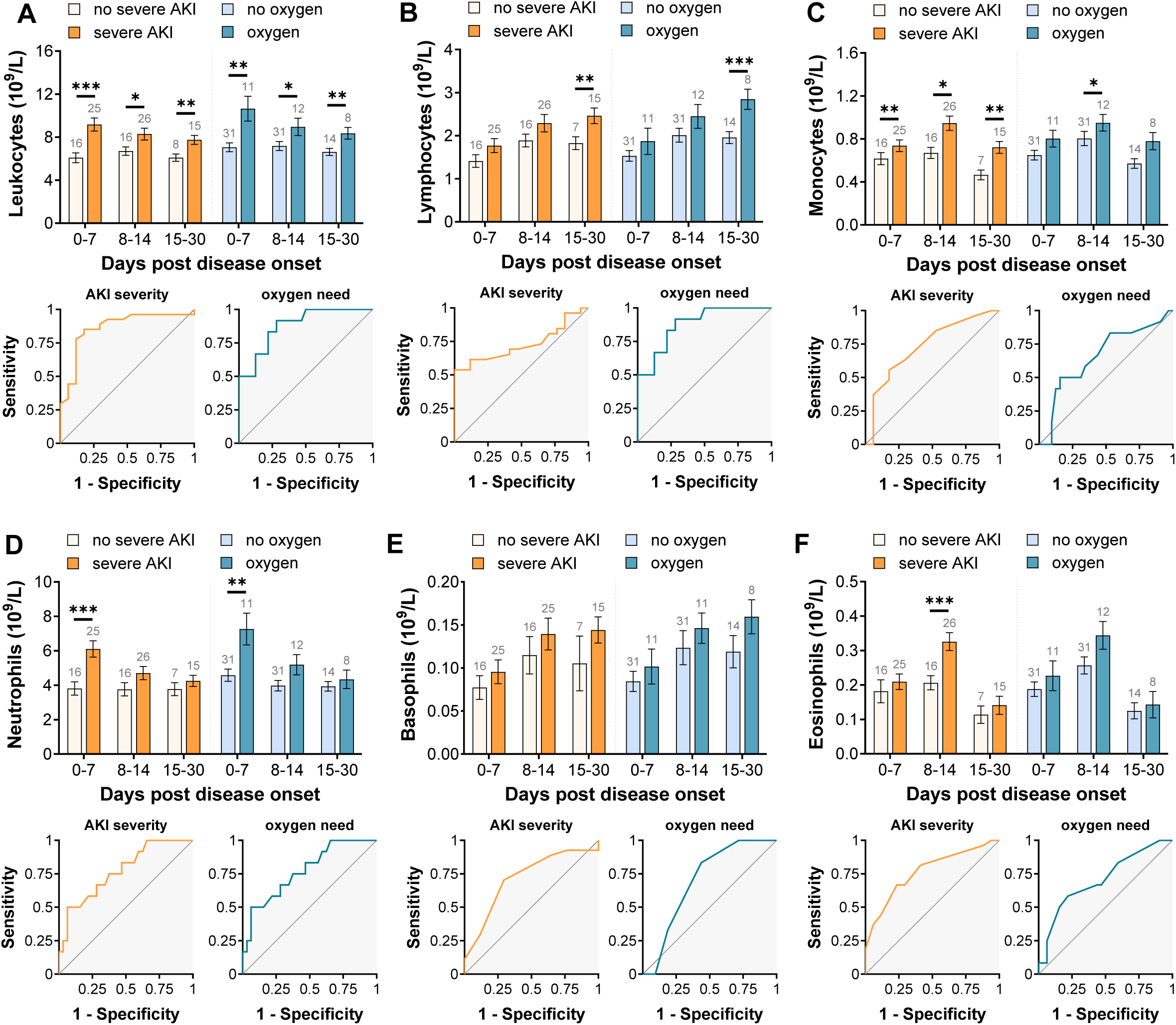
Immune cells levels in plasma for HFRS patients and association with acute severe injury or need of oxygen. HFRS patients were stratified into two groups of severe AKI (stage 2/3) or not (AKI stage 0/1) according to KDIGO criteria or in two groups need of oxygen or no oxygen. The estimated mean and standard error for each group were calculated using generalized estimating equation (GEE) and adjusted for sex and age. Receiver operating characteristics curve (ROC) analysis were performed using maximum values of eGLX markers and minimal values of albumin. The timeline kinetics and ROC curves are shown for leukocytes **(A)**, lymphocytes **(B)**, monocytes **(C)**, neutrophils **(D)**, basophils **(E)** and eosinophils **(F).** Significant differences within the same time point between marker levels are indicated by asterisks (*** p < 0.001; **p < 0.01; * p < 0.05). AKI acute kidney injury;

## Discussion

In this study, we demonstrated a significant association between eGLX degradation and renal and pulmonary dysfunction in HFRS patients. Our *in vitro* experiments further substantiated these observations, where we showed that MMP-9 and heparanase can degrade eGLX components. Importantly, albumin at physiological concentrations mitigated this degradation, preserving the integrity of the eGLX components. In endothelial cell model, MMP-9 disrupted the endothelial barrier, as evidenced by decreased impedance measurements and reduced levels of SDC-1 and HS at cell surface. However, albumin supplementation effectively attenuated this disruption, highlighting its protective role. Our findings suggest that eGLX degradation contributes to the pathogenesis of HFRS and may serve as a potential therapeutic target.

Our previous study was the first to demonstrate an association between elevated plasma levels of SDC-1 with Orthohantaviral disease severity, although the cohort was limited to 19 individuals^3^. Our findings were later confirmed by a study that showed that SDC-1 could predict severe AKI in patients infected with Hantaan virus, another Orthohantavirus^4^. In the present study, we extended our analysis to include several additional eGLX degradation markers: proteoglycans (SDC-1 and sTM), GAGs (HS), and plasma proteins (albumin). The degradation of the eGLX compromises the endothelial barrier function of kidney glomerular capillaries, increasing permeability and allowing proteins like albumin to leak into the interstitial space and into urine, contributing to hypoalbuminemia and proteinuria^23^. Consistent with findings from a Finnish cohort of 133 PUUV-infected patients, we observed transient, “flash-like” albuminuria during the early phase of the disease, supporting the role of eGLX disruption in the pathogenesis of HFRS-related renal dysfunction^24^. Moreover, eGLX degradation exposes endothelial cells to circulating inflammatory mediators and immune cells, promoting endothelial activation, inflammation, and microvascular thrombosis—all of which are potential mechanistic contributors to AKI^25^. By highlighting the degradation of multiple eGLX components and their association with AKI severity, our study provides further evidence that eGLX damage plays a critical role in the renal pathophysiology of HFRS.

Though renal injury is a central component of HFRS, pulmonary involvement is prominent in HFRS patients^26–30^. A previous study of HFRS patients showed pulmonary edema in nearly half of the patients as detected by computed tomography (CT) scans^27^. Our study identified 27% of patients in our cohort that required supplemental oxygen therapy, corroborating the involvement of pulmonary dysfunction in HFRS. Furthermore, our study showed a significant association between eGLX component levels and the need for oxygen supplementation. A potential pathogenic mechanism linking pulmonary eGLX degradation to oxygen need is the increased vascular leakage following eGLX degradation leading to leakage of plasma constituents into the alveolar spaces and interstitial tissue. This results in pulmonary edema for HFRS patients ^27^, that in turn impairs gas exchange by reducing the diffusion capacity of oxygen across the alveolar-capillary membrane, thereby causing hypoxemia and necessitating supplemental oxygen. Similar mechanisms including eGLX degradation have been observed in other lung injuries. Schmidt et al.^31^ demonstrated that during sepsis-induced lung injury, the eGLX undergoes significant degradation, evidenced by increased shedding of HS and decreased eGLX thickness. Pulmonary eGLX degradation has also been reported in acute respiratory distress syndrome (ARDS) caused by influenza virus infection^32^, contributing to increased vascular permeability and respiratory failure. These parallels suggest that eGLX degradation plays a critical role in the development of pulmonary dysfunction during Orthohantavirus infection.

Our *in vitro* studies identified a potential sheddase, MMP9, causing eGLX degradation during HFRS and increased vascular permeability which was abrogated by physiological levels of albumin. MMP9 has previously been shown to cause eGLX degradation in *in vitro* models^33,34^, an *ex vivo* renal model^35^ and an *in vivo* model of ARDS^36^. Furthermore, MMP9 released from Andes virus, an Orthohantavirus, infected dendritic cells caused vascular permeability^37^, supporting our findings and implicating MMP9 in Orthohantaviral disease pathogenesis. Patient levels of MMP9 were associated with renal and pulmonary dysfunction in our study, adding a clinical perspective to these findings. Though we did not find any significant changes in plasma levels of heparanase in our study, a previous study implicated this sheddase in AKI during HFRS^5^. In a previous study, HS removal from glomerular endothelial cells led to increased permeability for albumin^23^. We did not observe this in our *in vitro* studies; however, we identify albumin at physiological levels as protective against heparanase-mediated HS degradation. Potential sources of sheddases (MMP9 and heparanase) could be leukocytes, particularly monocytes and neutrophils, as the levels of these were highly correlated with disease outcome. Studies supporting these findings come from the Orthohantaviral-induced release of MMP9 from infected monocyte-derived dendritic cells^37^ and neutrophil release of heparanase leading to pulmonary damage in an *in vivo* model ^31^.

One of the strengths of our study is the analysis of consecutive patient samples using GEE, which allowed us to adjust for potential confounders such as sex and age. This statistical approach enhances the reliability of our findings by accounting for intra-individual correlations over time and providing robust standard errors. We also employed a comprehensive assessment of eGLX degradation by measuring multiple biomarkers that reflect its complex multilayered structure. Specifically, we quantified proteoglycans (SDC-1 and sTM), GAGs (HS), and plasma protein (albumin). This multifaceted approach provides a more detailed understanding of eGLX shedding and its association with disease severity in HFRS.

Despite these strengths, our study has limitations. The relatively small cohort size of 44 patients may limit the generalizability of our results to the broader population of HFRS patients. This sample size may not capture the full spectrum of disease presentations and could affect the statistical power of our analyses. Larger multicenter studies are warranted to validate our findings and to further elucidate the mechanisms underlying eGLX degradation in the pathogenesis of HFRS.

In conclusion, elevated levels of eGLX degradation markers are associated with severe AKI and the need for oxygen supplementation in HFRS patients. Although the exact mechanisms underlying eGLX shedding are not yet fully understood, our findings highlight the eGLX as a potential therapeutic target for improving outcomes in HFRS.

## Data Availability

All data produced in the present study can be made available as anonymized data upon reasonable request to the authors

## Acknowledgements

This study was funded by Region Västerbotten (Central ALF-funding: RV-836351, RV-967545; Base-unit ALF: RV-939769, RV-967783, RV-982300), Åke Wiberǵs Foundation (M18-0031), Molecular Infection Medicine Sweden (MIMS) Clinical Research Fellowship, Heart-Lung Foundation (20220179), Kempe Foundations (SMK21-0014). All of this funding was received by AMFC. We also would like to thank the Knut and Wallenberg Foundation (Wallenberg Centre for Molecular Medicine) for their support. We acknowledge Biochemical Macromolecular Characterization Umeå at Umeå university for support and access to the QCM-D instrument.

## Declaration of Interest

The authors declare no competing interests

## Declaration of generative AI and AI-assisted technologies in the writing process

During the preparation of this work the author(s) used ChatGPT to improve readability and language of the manuscript. After using this tool/service, the authors reviewed and edited the content as needed and take full responsibility for the content of the published article.

## Notes

### Competing Interest Statement

The authors have declared no competing interest.

### Funding Statement

This study was funded by Region Vaesterbotten (Central ALF-funding: RV-836351, RV-967545; Base-unit ALF: RV-939769, RV-967783, RV-982300), Ake Wibergs Foundation (M18-0031), Molecular Infection Medicine Sweden (MIMS) Clinical Research Fellowship, Heart-Lung Foundation (20220179), Kempe Foundations (SMK21-0014). All of this funding was received by AMFC. We also would like to thank the Knut and Wallenberg Foundation (Wallenberg Centre for Molecular Medicine) for their support. We acknowledge Biochemical Macromolecular Characterization Umea at Umea university for support and access to the QCM-D instrument.

### Author Declarations

The Swedish Ethical Review Authority gave ethical approval for this work.

