## Supplementary Figures for "Matrix Metalloproteinase-9 Mediates Endothelial Glycocalyx Degradation and Correlates with Severity of Hemorrhagic Fever with Renal Syndrome"

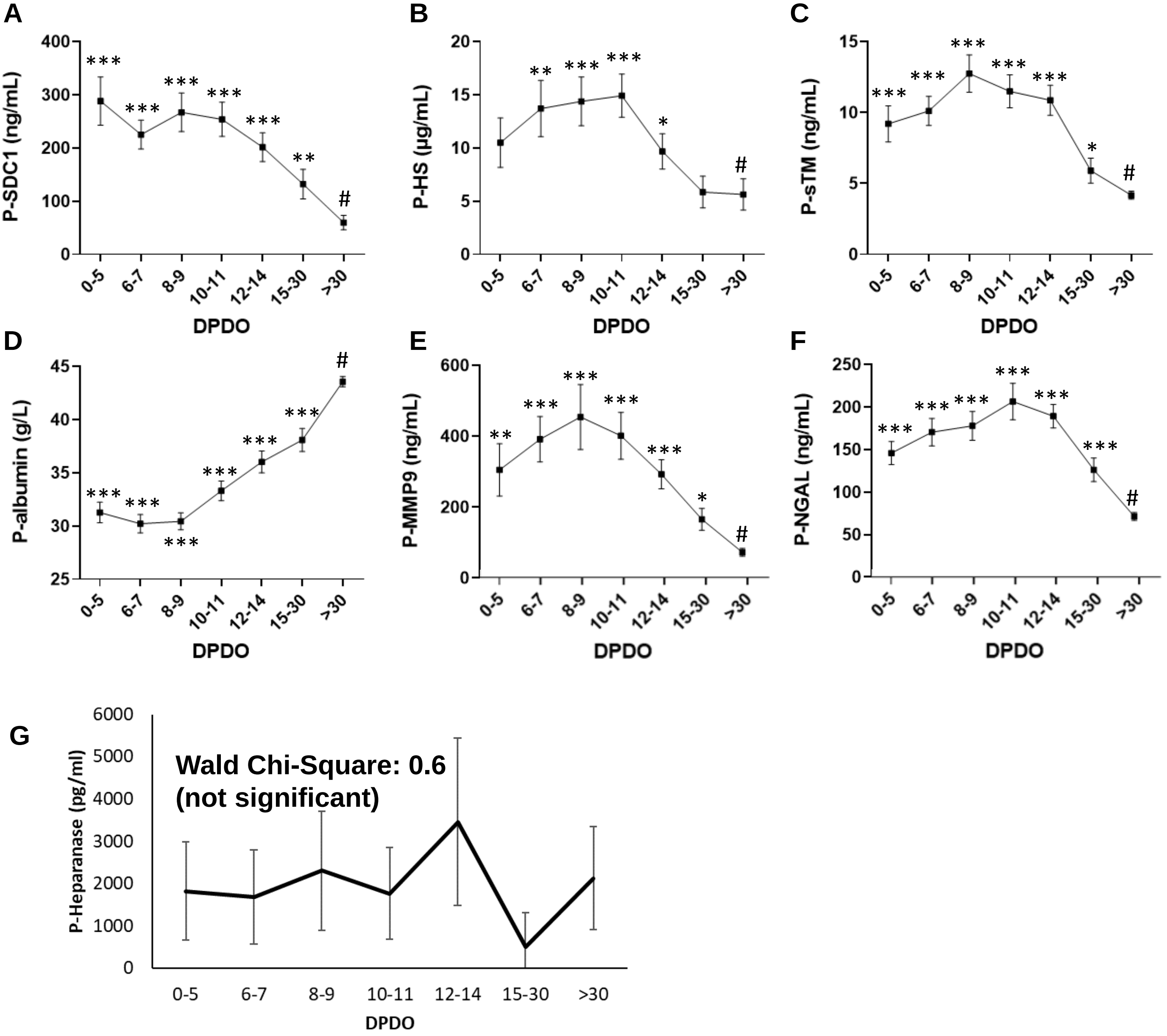


**Supplementary Figure 1: Kinetic of Endothelial glycocalyx degradation marker in HFRS patients in plasma.** The timeline kinetics are shown for syndecan-1 **(A)**, heparan sulfate **(B)**, soluble thrombomodulin **(C)**, albumin **(D)** MMP9 **(E)** NGAL **(F)** and heparanase **(G).** Significant differences between time points and convalescent point (#) are indicated by asterisks (*** $p < 0.001$; **$p < 0.01$; * $p < 0.05$).

HS heparan sulfate; SDC-1 syndecan-1; P plasma; sTM soluble thrombomodulin; MMP9 matrix metalloproteinase 9 ; NGAL neutrophil gelatinase-associated lipocalin.

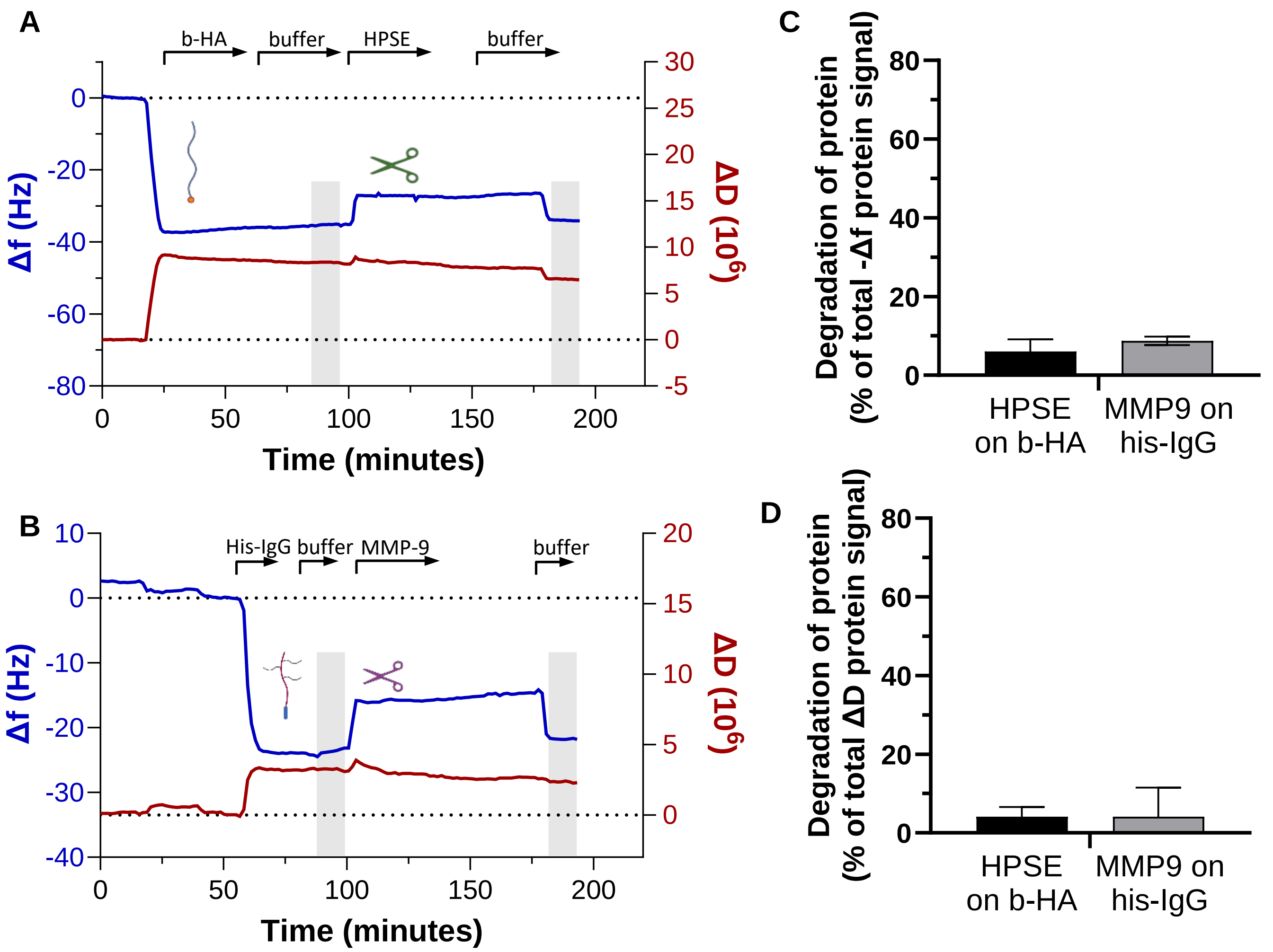


**Supplementary Figure 2: HPSE and MMP-9 specificity control using QCM-D. (A)** B-HA and **(B)** His-tagged IgG protein was absorbed on a synthetic supported lipid bilayer membrane composed by POPC-DOPE (95:5) lipids and streptavidin, plus Tris-NTA linker for the immobilization if his-tagged IgG. HPSE at 250 ng/mL and MMP-9 at 500 ng/mL were injected and their activity on b-HA or his-tagged IgG respectively, were followed. Frequency (Δf) and dissipation (ΔD) signals were compared before and after enzyme injection in buffer (grey rectangle areas) to assess the activity of the enzyme on the proteins and expressed as percentage reduction of -Δf and ΔD signal reduction for b-HA (**C**) and his-tagged IgG protein (**D**).

QCM-D quartz crystal microbalance with dissipation; b-HA biotinylated hyaluronan; IgG immunoglobulin G; MMP-9 matrix metalloproteinase 9; HPSE human heparanase.

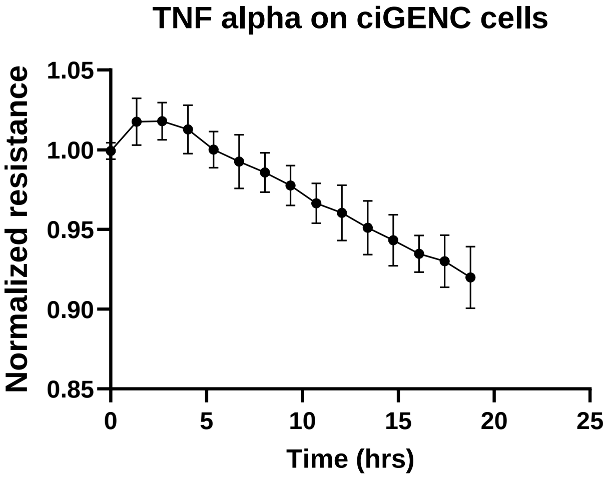


**Supplementary Figure 3: TNF-alpha effect on ciGENC cells.** Tumor necrosis factor-alpha (TNF-α; 50 ng/mL; was used as a positive control for barrier disruption on ciGENC cells. Endothelial barrier resistance was followed by impedance measurement (ECIS, APBiophysic).
